# The negative symptoms of schizophrenia: lessons from a precision nomothetic psychiatry approach

**DOI:** 10.1101/2022.05.26.22275663

**Authors:** Michael Maes

**Author notes:** **Corresponding author:** Prof. Dr. Michael Maes, M.D., Ph.D., Department of Psychiatry, Faculty of Medicine, Chulalongkorn University, Bangkok, 10330, Thailand, Michael Maes - Google Scholar.

## Abstract

The present study aims to explain how to use the precision nomothetic approach to analyze the interconnections between the negative symptoms, cognitive dysfunctions and biomarkers of schizophrenia. We review our data obtained in different study groups of patients with (deficit) schizophrenia and show, using examples extracted from these studies, how Partial Least Squares (PLS) path analysis should be used to examine these complex associations. PLS path analysis combines factor and multiple regression analysis in mediated models. We show that a single latent trait can be extracted from negative symptom domains and psychosis, hostility, excitation, mannerism, formal thought disorders and psychomotor retardation (PHEMFP). Both the negative and PHEMFP concepts miss discriminant validity whilst a common latent construct may be extracted from the 6 negative and 6 PHEMFP subdomains, dubbed overall severity of schizophrenia (OSOS). A common latent factor may be extracted from neurocognitive test scores including executive functions, and semantic and episodic memory dubbed the general cognitive decline (G-CoDe) index. PLS analysis shows that the effects of neuroimmunotoxic pathways on OSOS are partly mediated by the G-CoDe and indicate that those pathways have also direct effects on OSOS. We explain that the intercorrelations between those features should be assessed in an unrestricted study group combining patients and controls. Moreover, further bifactorial factor analysis with the restricted schizophrenia group may disclose illness-specific covariations among the features. Machine learning discovered a new schizophrenia phenotype characterized by increased severity of AOPs, G-CoDe, and OSOS, dubbed “major neurocognitive psychosis”.

## Introduction

In 2017, an estimated 1.1 million new cases of schizophrenia were reported, with a total of 20 million cases reported worldwide in 2019. ^1,2^ Between 0.3 percent to 0.7 percent of all individuals are diagnosed with schizophrenia. ^1-3^ About half of them will improve significantly over time, with no more relapses, and a small percentage will recover entirely, while the other half will be disabled for the rest of their lives. ^3-5^ People with schizophrenia frequently have long-term unemployment, poverty, and homelessness, they have a higher suicide risk and more physical health issues than the general population, resulting in a 20-year drop in life expectancy on average. ^6,7^

Dr. Emil Kraepelin published the first o0066ficial description of schizophrenia as a mental disorder in 1887. He coined the label “dementia praecox” or “early dementia” to characterize the symptoms that are now recognized as schizophrenia. The cornerstone of Kraepelin’s dementia praecox was a general “weakening” of mental processes that resulted in a “defect” coexisting with “productive” or “florid” symptoms. ^8-13^ In 1911, Eugen Bleuler coined the term “schizophrenia” a combination of the Greek terms schizo (split) and phrene (mind) to describe the mental disorder and fragmented thinking that affect people with schizophrenia. Bleuler considered schizophrenia to be a psycho-organic disorder comprising two symptom clusters, namely a core with four primary symptoms, namely disordered associations, autistic behavior and thinking, aberrant emotion, and ambivalence, and accessory symptoms including hallucinations, delusions, social disengagement, and reduced desire. ^14-16^

Kurt Schneider made the next significant advance in 1959 when he enumerated his ‘first rank’ characteristics of the disorder including “auditory hallucinations; thought withdrawal, insertion, and interruption; thought broadcasting; somatic hallucinations; delusional perception; feelings or actions as made or influenced by external agents”. ^16,17^ Since the 1970s, the labels “defect” and “productive symptoms” have mostly been replaced by the terms “negative” and “positive” symptoms. ^16^ Based on this new knowledge, new diagnostic criteria were constructed which are still used today by the International Classification of Diseases (ICD and the Diagnostic and Statistical Manual of Mental Disorders (DSM).

Crow presented a subclassification of schizophrenia depending on whether positive or negative symptoms predominate. “Type I” (positive) schizophrenia as defined by hallucinations, delusions, and formal thought disorder, with an underlying dopaminergic dysfunction, whereas “Type II” (negative) schizophrenia was defined by social withdrawal, loss of volition, affective flattening, and poverty of speech, all of which were presumed to be associated with structural brain abnormalities. ^18^ While positive symptoms may relate to dopaminergic aberrations in the mesolimbic circuits, negative symptoms were thought to be associated with dopaminergic aberrations in the mesocortical circuits. ^18^

Kirkpatrick et al. ^19,20^ suggested the definition of a subtype of schizophrenia defined by persistent “primary” negative symptoms that cannot be attributed to other psychiatric disorders. This clinical construct which was based on Kraepelin’s dementia praecox was dubbed “deficit schizophrenia” and was speculated to represent a unique “disease” within the schizophrenia spectrum. ^19,20,21^ When this negative symptom cluster is present throughout severe psychotic exacerbations and the more stable inter-episode stages of illness, it is referred to as deficit schizophrenia.

The NINH and NHS continue to categorize schizophrenia symptoms into three domains: positive and negative symptoms, as well as cognitive abnormalities. ^22,23^ Cognitive deficiencies associated with schizophrenia include impairments in working memory, executive functioning, fluency, list learning, attention and processing speed and those cognitive deficits contribute to lowered health-related quality of life (HR-QoL) and impairments in social functioning, including the ability to work, find work, live independently, and function normally. ^24-29^

Nonetheless, we discovered recently that a) negative and positive symptoms cannot be regarded as independent dimensions and that a latent vector underpins both positive and negative symptoms, dubbed the overall severity of schizophrenia (OSOS) latent vector; b) a common core of cognitive deficits underpins dysfunctions in episodic, semantic, and working memory, executive functions, and attention, dubbed the generalized cognitive decline (G-CoDe), c) the negative, positive and cognitive symptoms are strongly associated with neuroimmunotoxic pathways including increased levels of neurotoxic cytokines, chemokines, and oxidative and nitrosative stress compounds; and d) increased severity of OSOS, G-CoDe and neuroimmunotoxic pathways shape a new endophenotype class, namely major neurocognitive psychosis (MNP), largely overlapping with deficit schizophrenia. ^5,30-33^ Such data show that the common approach of examining the correlations between positive and negative symptoms, cognitive dysfunctions and biomarkers is inadequate and that these associations should be examined using the new precision nomothetic approach combining factor analysis and multiple regression analysis in Partial Least Squares (PLS) analysis. ^33,34^

### Aims of the study

The present study aims to explain how to use the precision nomothetic psychiatry approach to analyze the interconnections between positive and negative symptoms, cognitive dysfunctions and biomarkers in schizophrenia. Toward this end, we review our data obtained in different study samples of patients with schizophrenia and deficit schizophrenia and using examples extracted from these studies we show how PLS path analysis should be used to examine these complex associations. We will review how to use PLS and factor analysis to examine whether a common core underpins positive and negative symptoms and cognitive dysfunctions, and to examine how biomarkers and cognitive deficits predict the symptomatome of schizophrenia.

### PLS-path and factor analysis

PLS modeling allows for (a) the development of causal models that link causal factors including genetic and environmental factors (dubbed the “causome”) to adverse outcome pathways (AOPs), cognitive dysfunctions (dubbed the “cognitome”) and schizophrenia symptoms (dubbed the “symptomatome”); (b) the development and inclusion in the model of latent vectors based on unobservable variables, such as positive and negative symptom constructs; and (c) the delineation of mediation effects, such as the effects of AOPs on the symptomatome which are mediated by the cognitome. As input variables, different single indicators (age, gender, genomic data) and latent vectors derived from a set of highly connected indicators (e.g., a set of interconnected biomarkers or cognitive test results) may be used to predict the final outcome variable, namely the symptomatome. ^33,34^

The most prominent feature of PLS ^35,36^ is that linear composites of observed variables are constructed which act as proxies for latent variables that are difficult to measure directly, such as the ‘symptomatome”. ^33,34^ A latent construct could be thought of as something derived from empirical data that allows for empirical testing of the underlying constructs. ^33,34,37^ However, the model associations among the constructed latent vectors and single indicators can only be meaningfully evaluated if the PLS model’s construct validity is demonstrated. Therefore, the concept validity of the PLS model should be examined, including criterion validity, convergent validity, and discriminant validity. ^34,38^ Only when the model construct validity check meets predefined quality standards is a complete PLS analysis with 5000 or more bootstrap samples performed. The most critical conditions are that the model’s overall quality is acceptable, as indicated by a Standardized Root Mean Squared Residual (SRMR) <0.8, that the latent vectors demonstrate adequate reliability validity, as indicated by appropriate composite reliability >0.8, rho A > 0.8, Cronbach alpha > 0.7, adequate convergence as indicated by an average variance extracted > 0.5 and high factor loadings, namely > 0.6 at p < 0.001, and adequate prediction performance. The latter may be evaluated using PLSpredict and a tenfold cross-validation technique. ^33-35^ Moreover, Confirmatory Tetrad Analysis (CTA) should be conducted to confirm that the reflective models are not misspecified. Compositional invariance can be investigated using Predicted-Oriented Segmentation analysis and Measurement Invariance Assessment.

Furthermore, the produced latent vectors and indicators must have appropriate discriminant validity, which refers to how independent various latent components are different from one another. Discriminant validity assures that a construct is empirically unique and represents a reality that is not captured by other measures in the PLS model. ^36^ Checking for discriminant validity is a way to assure that the model’s latent vectors measure what they are supposed to measure. ^38^ Failure to disclose discriminant validity issues might lead to skewed structural parameter estimations and incorrect inferences regarding the associations between constructs. If the constructs do not have enough discriminant validity, they can be combined into a more generic construct that replaces the problematic constructs in the model, and researchers should re-evaluate the newly formed construct’s discriminant validity with all opposing constructs. ^38^

The Fornell-Larcker criterion, cross-loadings, and the Heterotrait-Monotrait (HTMT) ratio are three ways of determining discriminant validity. ^38^ The Fornell-Larcker criterion is used to assess the degree of shared variance between the model’s latent variables by comparing a construct’s AVE to its shared variance with other constructs, whereby its square root AVE (SRAVE) should exceed the correlations between the constructs. When each measured item has a poor association with every other construct except those to which it is conceptually related, discriminant validity is demonstrated. ^39^ A large correlation between items belonging to the same construct and a low correlation between items belonging to distinct constructs is necessary to demonstrate discriminant validity at the item level. Calculating cross-loadings, commonly known as “item-level discriminant validity,” is the second approach for evaluating discriminant validity. Each indicator loading should be bigger than all its cross-loadings, or the construct in question will be unable to distinguish between the construct it was designed to assess and another. ^40^ Double-loaders (variables that load heavily on two different factors) are a sign of poor discrimination validity. The most accurate method for measuring discriminant validity is perhaps the HTMT ratio ^38^ whereby an HTMT ratio > 0.85 ^41^ or > 0.9 ^42^ suggests a lack of discriminant ability.

Once the construct validity has been established, a full PLS path analysis can be performed using 5000 or more bootstrap samples, allowing for the computation of path coefficients with exact p-values, as well as specific indirect (or mediated), total indirect (mediated) and total (direct and indirect) effects, all of which are used to determine the significance of the (mediated) paths. As a result, PLS path modeling is an effective approach that allows the construction of novel models of a complex illness such as schizophrenia described by causal links between the causome, AOPs, cognitome, and symptomatome. ^33,34^ The PLS analyses that we conducted on different Thai and Iraqi study samples showed that the classical concept of negative symptoms needs revision.

### First issue: the labels “negative and positive symptoms” are confusing

The current standard view of schizophrenia is that it is divided into positive and negative symptoms. ^16,18,22,23^ Negative symptoms like emotional flatness, avolition, alogia, and anhedonia, as well as positive symptoms like delusions, hallucinations, excitement, and disordered thinking, ^43^ separate schizophrenia from other mental illnesses. The patient’s loss of emotions (anhedonia), cognitive processes (logic thinking), and behaviors (social isolation) are classified as negative symptoms, ^16,43,44^ whereas positive symptoms are assumed to be new and maladaptive mental processes and behaviors that did not exist before the onset of schizophrenia and have developed as symptoms of the illness. ^5,16,22,23,44^ Positive symptoms are changes in thoughts and feelings that are “added on” to a person’s experiences, whereas negative symptoms are “losses” that are “taken away” or diminished (e.g., lower motivation or reduced intensity of emotion) (e.g., paranoia or hearing voices). ^44^

Initially, Maes et al. ^30,31,45,46^ distinguished between positive and negative symptoms using standard rating scales such as the Positive and Negative Syndrome Scale (PANSS) ^47^ and Scale for the Assessment of Negative Symptoms (SANS) ^43^ as this is the gold standard method when analyzing schizophrenia data. ^5^ Nonetheless, Maes et al. ^46^ thought it would be fascinating to divide positive symptoms into their subdomains, such as psychosis (hallucinations and delusions), hostility, excitement, and mannerism, just as they thought it would be fascinating to investigate the subdomains of negative symptoms, such as anhedonia, avolition, and flattening. The central idea was that different symptoms may have distinct associations with biomarkers and molecular pathways. ^46^ Consequently, we created composite scores expressing psychosis, hostility, excitation, and mannerism (PHEM) based on all relevant z-transformed psychomotor retardation item scores of the PANSS and the Brief Psychiatric Rating Scale (BPRS), ^48^ whilst the negative symptom categories of the SANS were deemed appropriate. ^24,25,46^ In addition, formal thought disorders (FTD) and psychomotor retardation (PMR) were identified as two additional symptom domains that significantly contribute to schizophrenia symptomatology. ^49,50^ FTD is marked by abnormalities in abstract and concrete cognition, including disorganized, illogical, and inadequate mental processes, as well as intrusions, fluid thought, and weakened connections. ^49^ PMR is characterized by deficits in gross and fine motor function, sluggish motor responses, and slow movements. ^50,51^

The computed PMR score was based on the summed z transformed PMR item scores ^50,51^ from the PANSS, BPRS and the Hamilton Depression Rating Scale (HDRS). ^52^ Importantly, Cambridge Neuropsychological Test Automated Battery (CANTAB) assessments of the Motor Screening Task (MOT), ^53^ a psychomotor function index, externally confirmed this PMR score. ^50^

Nonetheless, a primary concern was that certain symptom domains, such as FTD and PMR, could not be properly classified as positive or negative. It is unclear, for instance, whether PMR is “added on” to a person’s normal motor behavior (a new motor dimension) or should be considered a “loss” (loss of movements). Similarly, the status of FTD is ambiguous: is it a novel thought process (the onset of formal cognitive disorders) or a loss of functions (loss of thought processes)? However, the same concerns can be asked regarding negative symptoms: are anhedonia and avolition, for example, really losses, or are they new “sensations” added to human experiences (“loss of feelings” is also a feeling)? And what about delusions? Do they add to the way people normally think or do they show a loss of normal human thought processes?

### A second issue: schizophrenia and range restriction

A second issue is that researchers, including Maes et al., ^45^ computed the associations between biomarkers and rating scale scores of positive and negative symptoms in schizophrenia study samples. Nevertheless, this study group is a restricted study sample of the population. Figure 1 (hypothetical data) shows the regression of severity of illness (the symptomatome severity or OSOS) on biomarker pathways (the APOs). This regression, for example, shows that around 63.6% of the variance in OSOS is explained by the severity of the AOPs when the regression is computed in controls (HC in this figure) and schizophrenia (SCZ in the figure) patients combined (F=44.79, df=2/51. P<0.001). However, if this regression is calculated in the restricted study sample of patients only, the association has less impact accounting for only 29.9% of the variance (F=7.69, df=2/36, p=0.002). Thus, when we restrict the range of OSOS (e.g. only patients are included) the correlation coefficient is reduced. Furthermore, if we narrow the range of OSOS even further (e.g., only patients with very high OSOS scores, SCZ2 in figure 1) the correlation may no longer be significant (F=0.23, df=2/18, p=0.796). This is a normal phenomenon resulting from restricting the range of the data. ^54,55^

**Figure 1.**
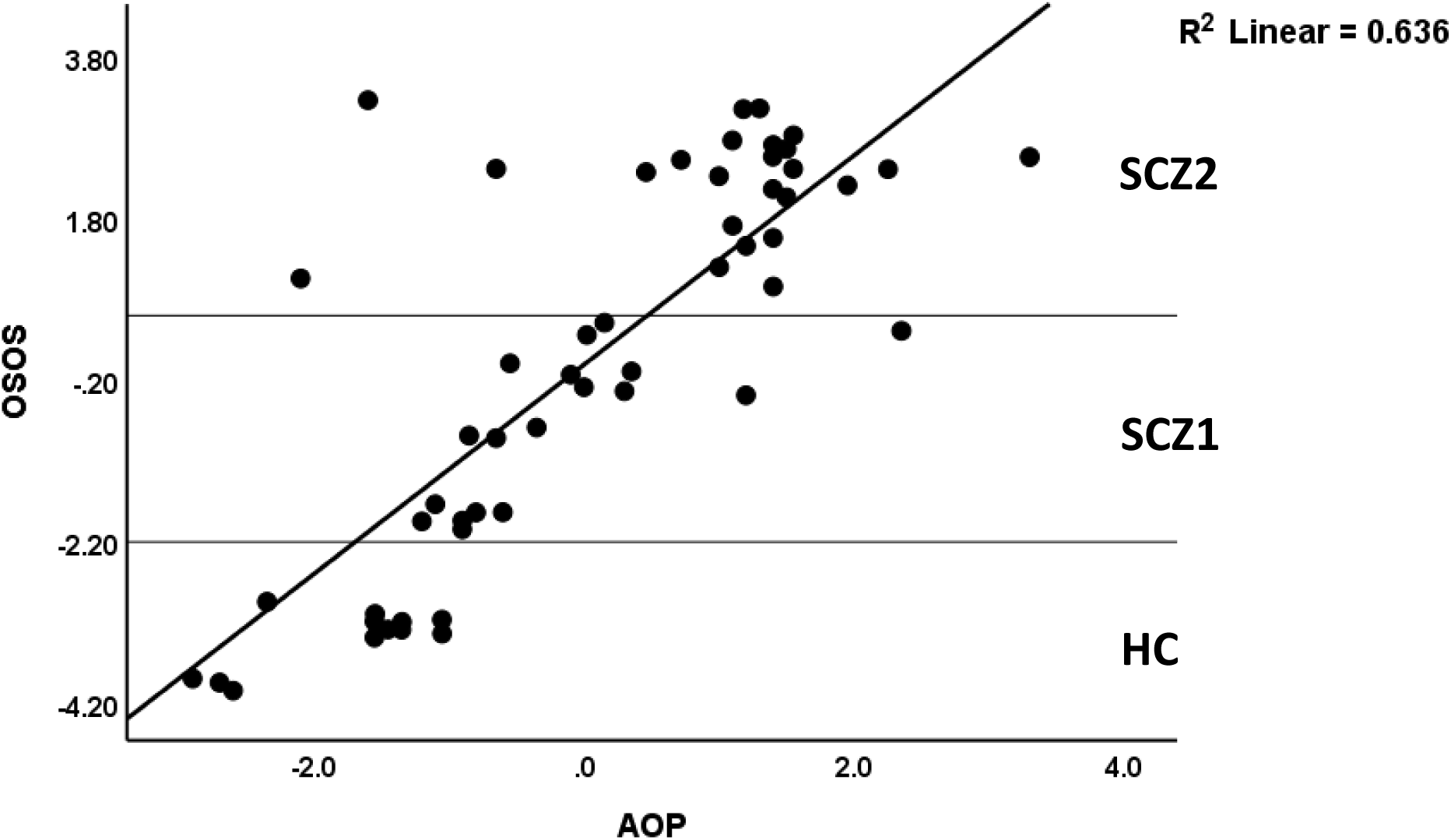
Regression analysis in (range restricted) study samples including healthy controls (HC) and different schizophrenia (SCZ) endophenotype classes (SCZ1 and SCZ2)

By analogy, it is common practice to compute differences in the biomarkers and severity of the symptomatome between controls and patients and to compute associations between biomarkers and severity in the restricted group of patients. ^5^ In fact, this is both illogical and incorrect. Thus, the regression of severity of illness on biomarkers is computed in a restricted sample, but the regression of the biomarkers on the classification (principle of ANOVA for regression) is carried out without any range restriction. ^5^ In the latter case, biomarkers are considered to be the dependent variable whereas, in reality, biomarkers are the explanatory variables that predict the severity of illness (dependent variable) which, in turn, is used to categorize the subjects into diagnostic classes. ^5^ Thus, the diagnosis of schizophrenia is a post-hoc man-made higher-order construct, which is the outcome variable but is always used incorrectly as an explanatory variable in analysthe is of variance or GLM analysis. It becomes even surreal when this outcome variable now employed as an explanatory variable shows very low reliability and validity, as reviewed previously. ^5^ As such, the majority, if not all, researchers use invalid higher-order constructs in incorrect statistical models (explanatory and dependent variables are switched) and use this category to range restrict when investigating covarions between features. Restricting sample variance artificially weakens existing correlations and generalizability and, consequently, correlation coefficients derived from an unrestricted sample should be corrected for range restriction using specific formulas to re-compute the actual correlation coefficient in an unrestricted sample. ^55,56^ Examining these associations in schizophrenia models, we have repeatedly shown that PLS-POS, a method to disclose heterogeneity in PLS path models, did never retrieve the diagnosis as a source of heterogeneity or segmentation.

### Third issue: PHEMFP and negative symptoms do not show discriminant validity

Figure 2 illustrates an example of a PLS model in which the effects of biomarkers on negative symptoms and PHEMFP symptoms are investigated. Given that we discovered strong associations among the 6 PHEMFP symptom domains and the negative symptom domains of the SANS, we first investigated whether it was possible to derive reliable latent vectors from the PHEMFP and negative symptom domains. This model is based on the data presented in Maes et al. ^51^ and is, therefore, based on real data collected from schizophrenia patients and controls.

**Figure 2.**
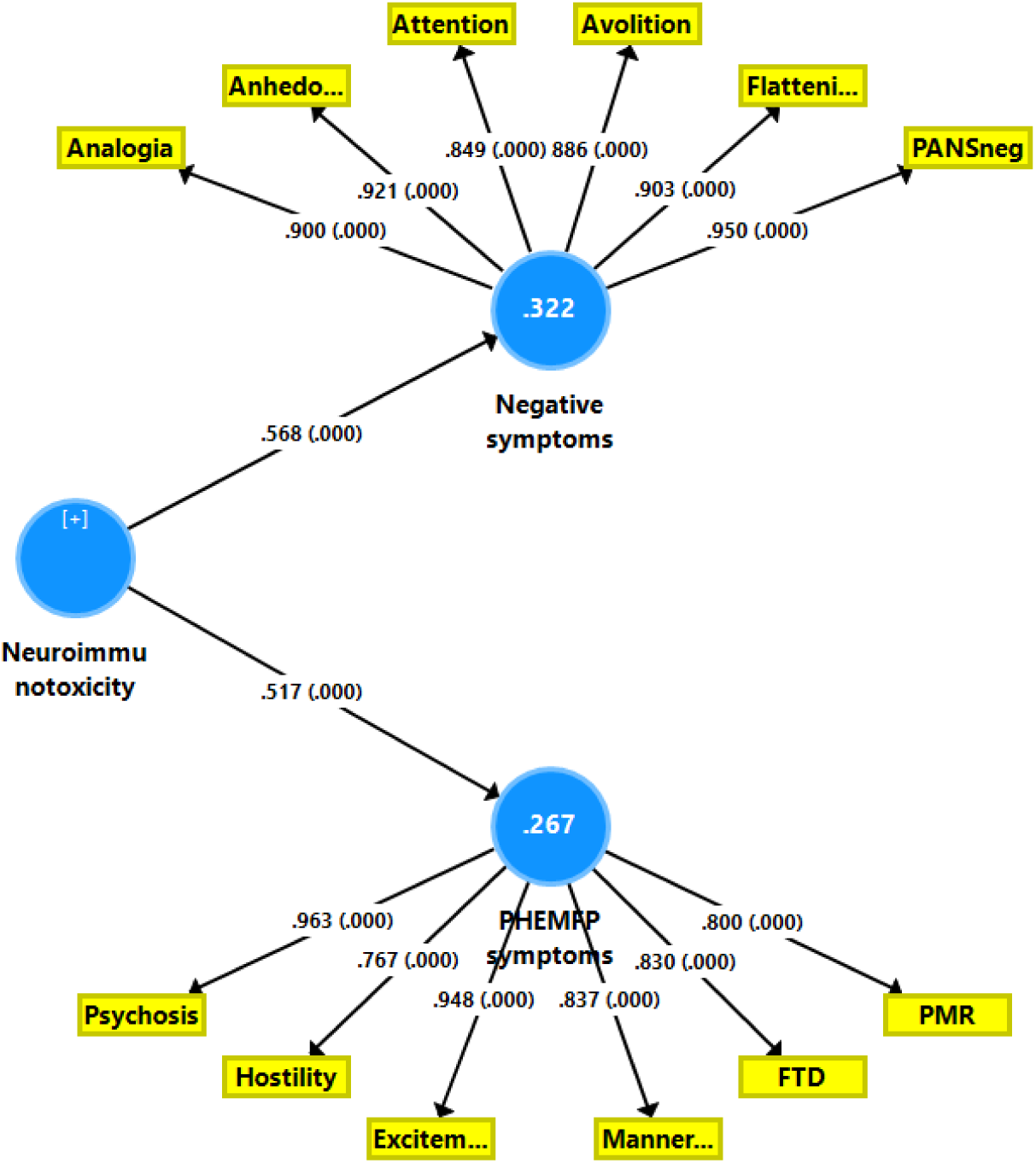
Results of Partial Least Squares (PLS) analysis. The neuroimmunotoxic adverse outcome pathways predict two different latent vectors extracted from either negative symptom domains (analogia, anhedonia, avolition, attention, flattening, PANNS negative scale score) or PHEMFP (psychosis, hostility, excitation, mannerism, formal thought disorders (FTD) and psychomotor retardation (PMR)). Shown are the loadings (with p values) of all indicators of the latent vectors and the path coefficients (with p values). Figures in the blue circles indicate explained variance. PANSneg: negative domain score of the Positive and Negative Syndrome Scale (PANNS)

Firstly, the data demonstrate that we were able to extract a reliable vector with adequate construct validity that underpins the negative symptom domains because a) the composite reliability (=0.963), rho A (=0.959), and Cronbach alpha (=0.954) values are more than adequate, demonstrating accurate internal consistency or composite reliability; b) the AVE value was 0.814 indicating sufficient convergent validity; c) all loadings on the latent vector are significant and > 0.849, and d) CTA confirms that the vector is not misspecified as a reflective model. As a result, the six negative subdomains are manifestations of this latent vector known as the “negative symptom domain.” In addition, the first latent vector extracted from the PHEMFP symptoms demonstrates a) adequate composite reliability (=0.945), rho A (=0.946), and Cronbach alpha (=0.929); b) adequate convergence with AVE of 0.741 and significant loadings all greater than 0.767; and c) CTA results confirming that this factor is not misspecified as a reflective model. Consequently, the PHEMFP symptom domains should also be viewed as manifestations of this latent single trait. In addition, PLS blindfolding revealed that the negative (0.253 and PHEMFP (0.187) latent vectors exhibit appropriate construct cross-validated redundancies, indicating that both constructs have predictive significance. Lastly, the model quality data are more than adequate, with SRMR = 0.041 whilst PLS predict demonstrates that the Q2 predict scores for all indicators are positive, indicating that they outperform the most naive benchmark and, therefore, that the model has significant predictive performance.

Based on this high-quality model data, we performed a complete PLS analysis using 5000 bootstrap samples to investigate the pathways from biomarkers to both positive and negative symptoms. To display a more parsimonious model, we entered the biomarkers as a single indicator, namely a z unit-based composite score computed as the sum of the z transformations of various cytokines + z chemokines + z oxidative stress biomarkers + z lipopolysaccharides + z leaky gut indicators + z BBB permeability indicators. ^33,57^ The final PLS model reveals that the combined effects of neuroimmunotoxic pathways account for 32,2% of the variance in the domain of negative symptoms and 26,7% of the variance in the domain of PHEMFP.

Nonetheless, this model has significant flaws because both latent vectors lack discriminatory power. First, the Fornel-Larcker criterion (see **Table 1**) demonstrated that the SRAVE of the PHEMFP construct was less than its correlation with the negative symptom subdomain. Second, the cross-loadings (**Table 2**) demonstrate that all negative and PHEMFP domain scores have a high cross-loadings on both factors and thus that all subdomains are double-loaders. All indicator loadings are indeed greater than their cross-loadings (except PMR), but all PHEMFP symptoms loaded highly on the negative latent vector (e.g., excitement loaded at 0.948 on the PHEM vector and 0.856 on the negative domain vector) and all negative subdomains loaded highly on the PHEMFP subdomain (e.g. avolition showed a loading of 0.886 on the negative vector and of 0.799 on the PHEMFP vector). Thirdly, the HTMT ratio between PHEMFP and the negative domain was 0.906, indicating a lack of discriminatory ability. We have rerun these tests of discriminant validity with the same six negative subdomains and the four PHEM subdomains (psychosis, hostility, excitation and mannerism). Similar results to those described in the preceding section indicate that these constructs lack sufficient discriminant validity with, for example, an HTMT ratio of 0.862. In such situations, it is recommended to reassign problematic indicators or combine problematic latent vectors. Since most of the indicators appear problematic, we looked into whether or not both vectors could be combined into a single overarching latent structure.

**Table 1.** Discriminat validity (Fornel-Larcker criterion) among features of schizophrenia, namely negative and PHEMFP (psychosis, hostility, excitation, mannerism, formal thought disorders, psychomotor retardation) symptom domains and neuroimmunotoxicity (NIT)

| Features | NIT | Negative domains | PHEMFP domains |
| --- | --- | --- | --- |
| NIT | 1.0 |  |  |
| Negative domains | 0.568 | 0.902 |  |
| PHEMFP domains | 0.517 | 0.871 | 0.861 |

**Table 2.** Discriminat validity (cross-loadings) among negative and PHEMFP (psychosis, hostility, excitation, mannerism, formal thought disorders, psychomotor retardation) symptom domains

| Symptom domains | Negative latent vector | PHEMFP latent vector |
| --- | --- | --- |
| Psychosis | 0.781 | 0.963 |
| Hostility | 0.605 | 0.767 |
| Excitation | 0.856 | 0.948 |
| Mannerism | 0.674 | 0.837 |
| Formal thought disorders | 0.652 | 0.830 |
| Psychomotor retardation | 0.846 | 0.800 |
| PANNS negative domain | 0.950 | 0.864 |
| Avolition (SANS) | 0.886 | 0.799 |
| Anhedonia (SANS) | 0.921 | 0.806 |
| Analogia (SANS) | 0.900 | 0.780 |
| Attention (SANS) | 0.849 | 0.698 |
| Flattening (SANS) | 0.903 | 0.752 |

**Figure 3** shows a new PLS model with the 6 negative and 6 PHEMFP subdomain scores combined. Surprisingly, one vector could be extracted from the 12 subdomains and this factor showed adequate construct validity with accurate composite reliability (=0.969), rho A (=0.969), Cronbach alpha (=0.964) and AVE (=0.723)values, and significant loadings (>0.707) that were all significant (p<0.001). Furthermore, CTA confirmed that the vector is not misspecified as a reflective model, whilst blindfolding showed an appropriate cross-validated redundancy (0.408) indicating that this latent vector has good predictive relevance. As such, we concluded that negative and PHEM symptoms lack sufficient discriminatory power and probably do not constitute distinct constructs. All negative and PHEM(PF) subdomains are manifestations of the same common latent trait.

**Figure 3.**
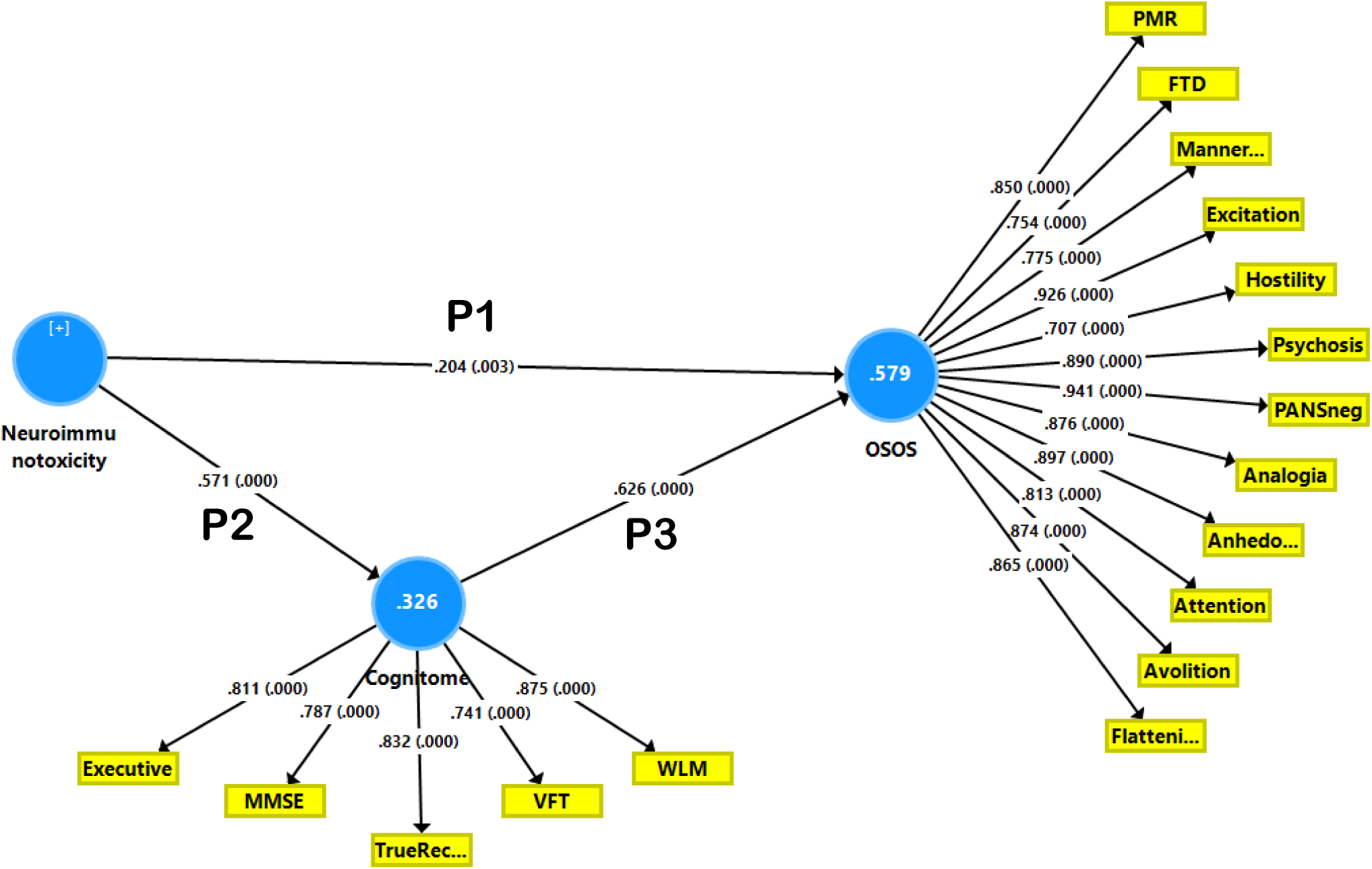
Results of Partial Least Squares (PLS) analysis. The neuroimmunotoxic adverse outcome pathways and a latent vector extracted from cognitive test scores (dubbed the cognitome) predict a latent vector extracted from negative (analogia, anhedonia, avolition, attention, flattening, PANNS negative scale score) and PHEMFP (psychosis, hostility, excitation, mannerism, formal thought disorders (FTD) and psychomotor retardation (PMR)) symptom domains. The cognitive test scores are: Mini Mental State Examination (MMSE), executive functions tests, verbal fluency test (VFT), Word List Memory (WLM) and True Recall Shown are the loadings (with p values) of all indicators on the latent vectors and the path coefficients (with p values). Figures in the blue circles indicate explained variance. P1: direct effects, P1 and P2 mediated (indirect) effects PANSneg: negative domain score of the Positive and Negative Syndrome Scale (PANNS)

Almulla et al., ^58^ using a study sample consisting of Iraqi schizophrenia patients and controls, examined the factor structure of the 6 PHEM symptoms, the total SANS score and the negative subscale of the PANNS using Exploratory Factor Analysis (EFA) including dimensionality tests which allow estimating the number of factors to be retained including Parallel Analysis (Optimal Implementation), the Schwartz’s Bayesian Information Criterion (BIC) and the Hull test. ^58^ Furthermore, these authors assessed closeness to unidimensionality utilizing unidimensional congruence (UNICO), explained common variance (ECV) and mean of item residual absolute loadings (MIREAL) whereby the data should be treated as essentially unidimensional when UNICO >0.95; ECV >0.85; and MIREAL <0.300. ^58^ The EFA results indicated that all subdomains loaded highly on the first factor (all > 0.660) while the dimensionality tests showed that only one factor should be retained. The closeness to unidimensionality tests indicated that the PHEMFP and negative symptoms should be treated as essentially unidimensional. ^58^ Moreover, model fit indices showed excellent model fit, good construct replicability, excellent performance across studies and good quality of the factor score estimates.

All in all, the above results of different studies ^33,58^ show that a single latent trait, which is essentially unidimensional, underpins the 6 PHEMFP and negative symptoms of schizophrenia and therefore that the latent variable score of these subdomains may serve as a validated and reproducible index of OSOS. ^51,58^ Our results indicate that OSOS can be reflectively quantified using PHEMFP and negative subdomain scores and that this reflective latent construct serves as the common denominator for the manifestations that are largely determined by OSOS. It follows that studies reporting on differential correlations between biomarkers and positive and negative symptoms are not very relevant, especially not when computed in the restricted study group of schizophrenia patients.

### Analyzing feature associations within the restricted schizophrenia group

In our studies described above, we also performed PLS or EFA using the same variables in the restricted study group of schizophrenia patients. ^51,58^ These results showed again that one factor albeit less significant may be extracted from the PHEMPF subdomain and negative PANSS and SANS scores, indicating that even in a restricted study sample, the same latent trait could be established. These findings again show that selecting only schizophrenia to compute correlations between OSOS and biomarkers restricts the range and thus may artificially weaken existing correlations, thereby hampering generalizability.

Nevertheless, we also analyzed our data using Bifactorial Direct Hierarchical EFA, which allows us to define a first generalized factor and additionally one or more single-group factors. ^51^ A bifactor model differs considerably from classical factor analysis with rotated solutions because a bifactor model method defines a general factor and subordinate or single group factors. The former is a broader factor which is the source of common variance running through all items in the EFA and the single group factor reflects the coherency among a subgroup of items not accounted for by the general factor. Maes et al. ^51^ detected that the PHEMFP and total SANS and PANSS negative symptom scores were best modeled by employing a bidimensional oblique solution consisting of a) a general factor (GF) reflecting OSOS; and b) a single-group factor (SGF) reflecting negative symptoms and PMR combined. Importantly, the same bifactorial exploratory solution with a general factor reflecting OSOS and a single group factor reflecting a subordinated factor of negative symptoms and PMR was detected in the restricted sample of schizophrenia patients. Moreover, the general and single group factor scores were differentially associated with biomarkers pathways indicating that both constructs are mediated by different albeit partially overlapping pathways. This shows that it is also important to compute the associations between biomarkers, OSOS and putative single group factors in the restricted study group of schizophrenia patients.

All in all, computing the associations between dependent symptomatome (e.g. OSOS and the single-group factor) and independent (causome, environmentome, AOPs, cognitome, connectome) variables in the unrestricted study group of controls and schizophrenia patients combined allows defining the covariation between those components all over the spectrum from controls to the schizophrenia spectrum including from mild to the most severe phenotypes. This approach should be complemented by the computation of the regression between the features in the restricted sample of schizophrenia patients to delineate schizophrenia-specific covariations among those variables.

Our findings that a unidimensional or bifactorial model is the best way to explain the symptomatome of schizophrenia contradicts the conventional gold standard, which holds that a two-dimensional construct consisting of positive and negative symptoms is the gold standard. **Figure 4** shows the covariation between OSOS and AOPs all over the spectrum from controls to schizophrenia patients and the more severe phenotypes as well as the association between the single-group factor and AOPs which together shape a distinct more severe phenotype characterized by increased OSOS and single-group factor scores and more severe AOPs (see below).

**Figure 4.**
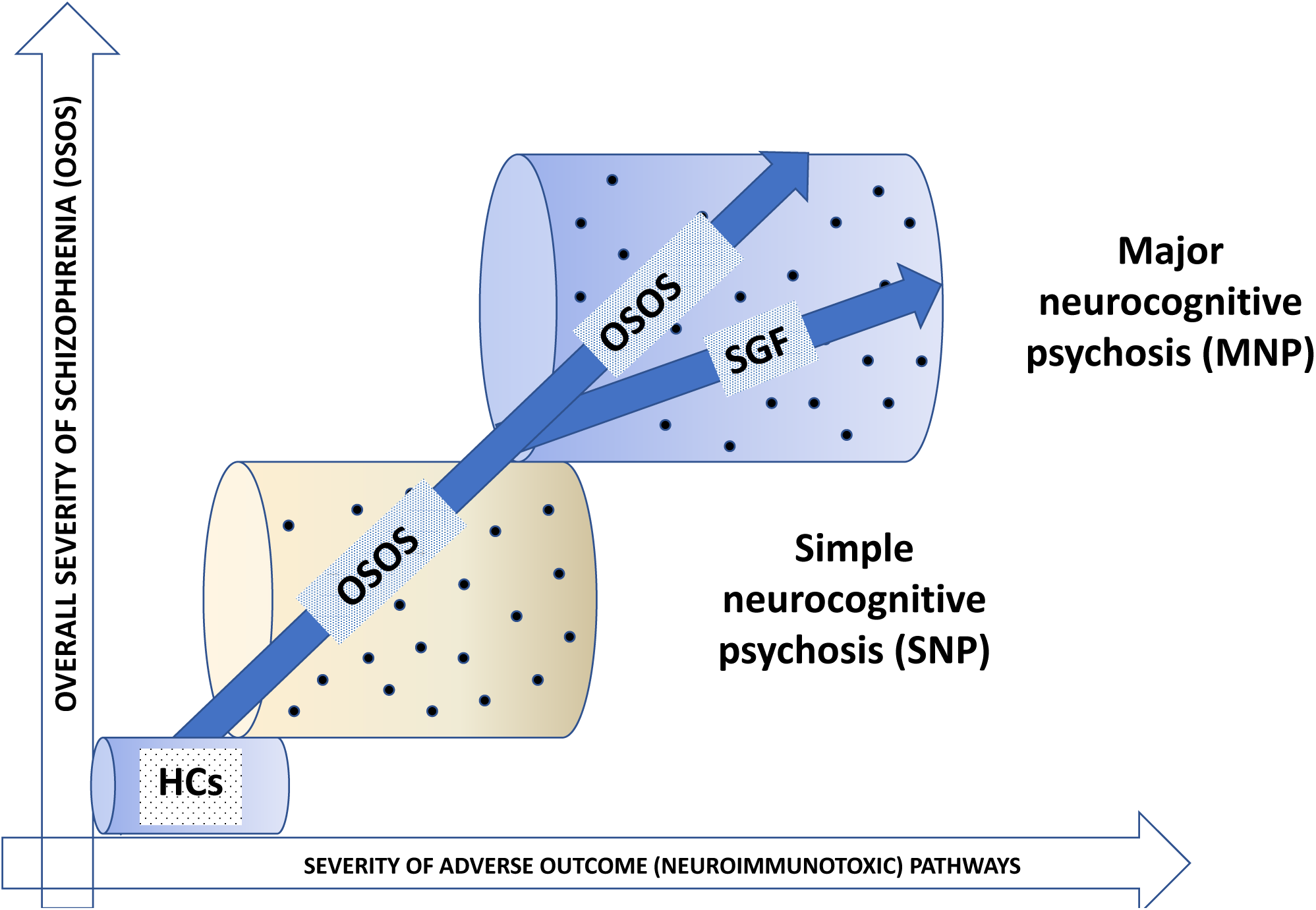
Position of patients belonging to qualitatively distinct schizophrenia endophenotype classes in a spectrum of increasing adverse outcome pathways and overall severity of schizophrenia from healthy controls (HCs) to simple neurocognitive psychosis (SNP) and the more severe endophenotype class MNP. SGF: single group factor consisting of negative symptom and psychomotor retardation Adapted from Maes et al., 2019

### The G-CoDe

We and others have previously discussed how deficits in executive functions, learning and working memory, as well as semantic and episodic memory, can result in the formation and recall of false memories, and thus may partially explain formal thought disorders, disorganized thought processes, paranoia, and other schizophrenia symptoms. ^24,29,59,60^ Cognitive impairments that reflect central neurocircuitry dysfunctions frequently precede the onset of acute psychotic episodes, suggesting that cognitive abnormalities contribute to the schizophrenia symptomatology. ^24,29^ For ultra high-risk individuals, verbal memory deficits and attentional deficits, for example, are predictive of psychotic symptoms. ^61,62^

Given that cognitive impairments may precede and explain at least a part of the schizophrenia symptomatology, we have incorporated cognitive impairments into our PLS models as shown in Figure 3 and allowed cognitive impairments to predict the symptomatome and AOPs to predict cognitive deficits and the symptomatome. As a result, a mediation model is developed in which cognitive deficits fully or partially mediate the effects of AOPs on the symptomatome. In our initial attempts to investigate such mediated effects, we separately analyzed cognitive disorders as measured by different probes of the Consortium to Establish a Registry for Alzheimer’s Disease (CERAD) ^63^ and the CANTAB.

Nevertheless, when examining the highly significant intercorrelations and cross-loadings between the cognitive test scores of CERAD and CANTAB, we combined different executive CANTAB test scores (spatial working memory, one-touch stockings of Cambridge and intra-extra dimensional set-shift) with the CERAD tests’ scores (verbal fluency, word list memory, recall and the Mini-Mental State Examination (MMSE) scores) in factor analyses. Figure 3 demonstrates that we were able to extract one latent vector from those multiple cognitive test scores that showed adequate psychometric properties, namely: a) adequate internal consistency with composite reliability of 0.905, rho A of 0.875, and Cronbach alpha of 0.869; b) adequate convergent validity with AVE = 0.657 and all loadings on the latent vector are > 0.741 at p<0.001; c) adequate construct cross-validated redundancy of 0.207, and d) confirmation that the vector is not misspecified as a reflective model. Impairments in semantic and episodic memory, recall, MMSE, and executive functions are thus all manifestations of a single trait that is the cause of these cognitive dysfunctions. As a result, we coined this latent vector “generalized cognitive decline” (G-CoDe) because the severity scores indicate gradual and more generalized deterioration of cognitive functions. ^24^ Importantly, those neurocognitive deficits in schizophrenia reflect dysfunctions in brain circuits, including “the prefronto-parietal, prefronto-temporal, prefronto-striato-thalamic, and dorsolateral prefrontal cortex, amygdala, and hippocampus.” ^24,59^

Figure 3 depicts the final PLS model, which has more than adequate model quality, with an SRMR of 0.043. We found that 57.9% of the variance in the symptomatome was explained by the regression on the G-CoDe and the AOPs, while 32.6% of the variance in the cognitome was explained by the regression on the AOPs. This PLS graph depicts the path coefficients (with exact p-value) of the causal relationships between the various indicators. This figure also displays the direct effects of AOPs on the symptomatome (P1), the indirect (or mediated) effects of AOPs on the symptomatome through the cognitome (P2. P3), and the total effect, namely P1 + (P2. P3). Significant specific indirect effects (t=5.70, p<0.001) of the AOPs on the symptomatome were mediated by the cognitome, which serves as a successful partial mediator (partly because there are also direct effects of the AOPs on the symptomatome). The total effects of the AOPs on OSOS are highly significant (t=8.55, p<0.001). As a result, a simple mediator model is constructed, indicating that the AOPs have both direct and indirect (or mediating) effects on the symptomatome. This is an example of complementary mediation in which the direct and indirect effects are complementary because they are both significant and point in the same direction. In addition, this is a ddistal-mediatedmodel because the path from the cognitome to the symptomatome is greater than the path from the AOPs to the cognitome. Importantly, such results demonstrate that neuroimmunotoxic AOPs cause aberrations in prefronto-parietal, prefronto-temporal, prefronto-striato-thalamic, and dorsolateral prefrontal cortical, amygdaloid, and hippocampal circuits, and that these circuits partially determine OSOS. In addition, the direct effects of AOPs on the symptomatome demonstrate that other pathways unrelated to the cognitome are also involved in the AOPs’ effects on OSOS. It should be emphasized that the Fornell-Larcker criterion revealed that the cognitome and OSOS have sufficient discriminant validity, although the HTMT was 0.850, indicating that discriminant validity may not be achieved. Nonetheless, the cross-loadings showed that each cognitome indicator loaded highly on OSOS and that each of the 12 OSOS indicators loaded highly on the cognitome factor. Therefore, we deemed it essential to determine whether a single latent trait could be extracted from the five cognitome and twelve OSOS domains. **Figure 5** demonstrates that one latent trait could be extracted from these 17 indicators with sufficient construct validity (all loadings > 0.610 at p<0.001, AVE=0.632, composite reliability=0.966, rho_A=0.967 and Cronbach alpha=0.962, construct cross-validated redundancy=0.215). In addition, CTA demonstrated that this construct was not incorrectly specified as a reflective model. This demonstrates that a single common core underlies the cognitive deficits, PHEMFP and negative symptoms of schizophrenia, indicating that objective neurocognitive deficits and the symptomatome are strongly intertwined indicators of the core of schizophrenia. Since the neurocognitive impairments measured in schizophrenia reflect aberrations in cortico-amygdala-hippocampal circuits, it follows that also OSOS is largely mediated by dysfunctions in these circuits. Moreover, PLS path analysis revealed that the regression on the AOPs explained 35.8 percent of the variance in this common core. Inferentially, the neuroimmunotoxic AOPs contribute to aberrations in the cortico-thalamo-hippocampal circuits leading to the cognitome and OSOS, whereby the pathway from AOPs to cognitome and OSOS is a key component of schizophrenia.

**Figure 5.**
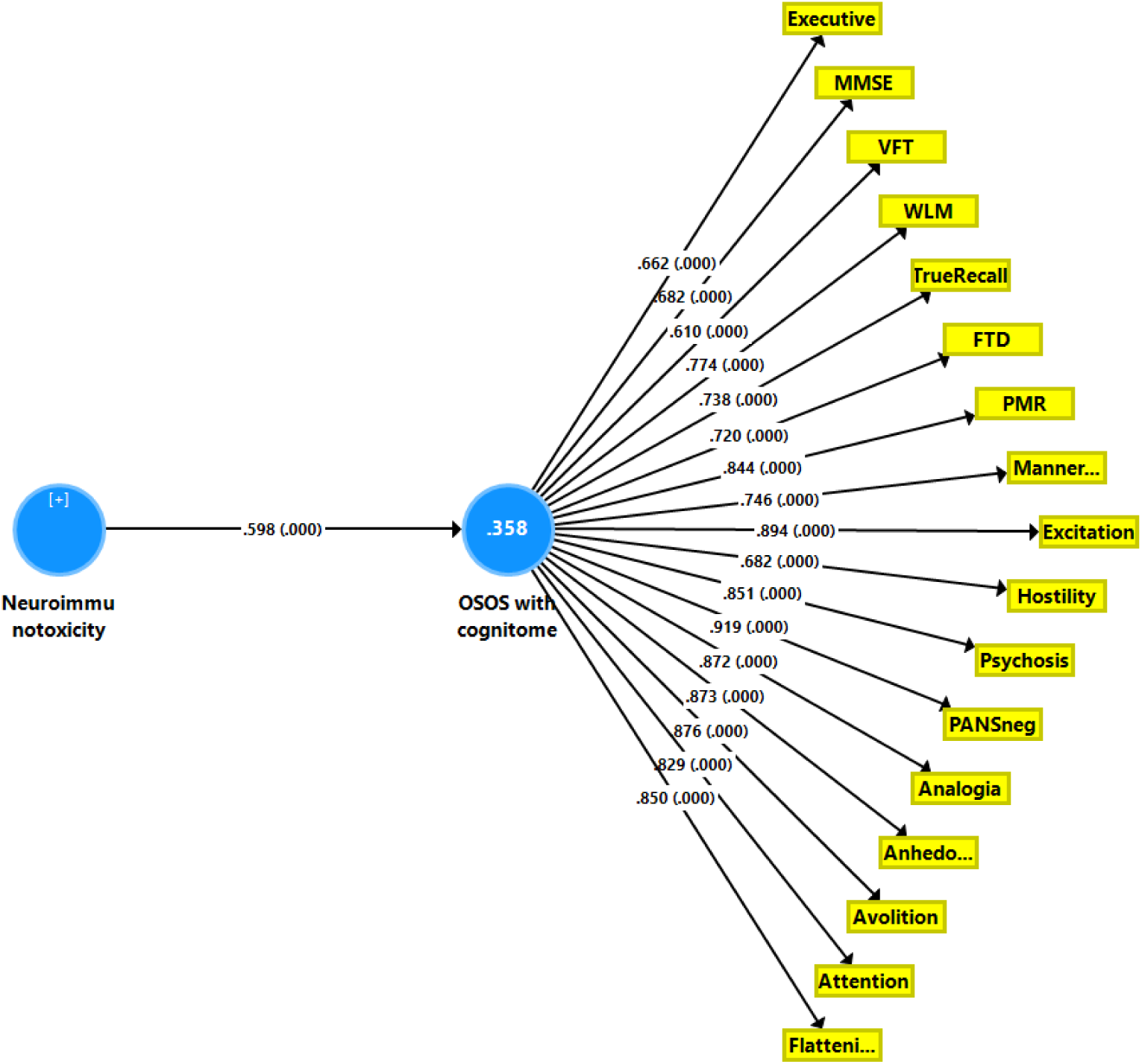
Results of Partial Least Squares (PLS) analysis. The neuroimmunotoxic adverse outcome pathways predict a latent vector extracted from cognitive test scores (Mini Mental State Examination (MMSE), executive functions tests, verbal fluency test (VFT), Word List Memory (WLM) and True Recall), negative symptom domains (analogia, anhedonia, avolition, attention, flattening, PANNS negative scale score) and PHEMFP symptom domains (psychosis, hostility, excitation, mannerism, formal thought disorders (FTD) and psychomotor retardation (PMR)). Shown are the loadings (with p values) of all indicators of the latent vector and the path coefficient (with p values). Figures in the blue circles indicate explained variance. PANSneg: negative domain score of the Positive and Negative Syndrome Scale (PANNS)

### Major neurocognitive psychosis and deficit schizophrenia

The results of the aforementioned PLS analyses are crucial for understanding the phenome of the newly discovered endophenotype classes of schizophrenia, namely “Major Neurocognitive Psychosis” and “Simple Neurocognitive Psychosis” (SNP). ^30,58,64^ Using unsupervised learning (cluster analysis) on Thai and Iraqi study groups with clinical, cognitive, and AOP data as input variables, a two-cluster solution was discovered with the separation of stable phase schizophrenia into two clinically and biologically meaningful clusters. ^30^ Using conventional statistical tests, we determined that MNP is distinguished from SNP and healthy controls not only by increased negative and PHEMFP subdomain scores, but also by increased affective symptoms, more cognitive deficits, and aberrations in a multitude of AOPs. ^30,58,64^ In addition, MNP is accompanied by highly specific abnormalities in protective AOPs (dubbed the “protectome”) including natural IgM-mediated autoimmune responses, antioxidant gene variants and diminished antioxidant defenses, which are absent in SNP and thus are distinguishing features of MNP. ^51^ Therefore, MNP is a clinically and biologically distinct group that can be differentiated from SNP. Furthermore, using a specific supervised learning technique that allows for the examination of whether a class is qualitatively distinct (i.e., soft independent modeling of class analogy or SIMCA), we were able to confirm that MNP is qualitatively distinct from SNP based on negative, PHEMFP and neurocognitive scores and AOPs as well, ^30,58,64^ indicating that MNP is validated as a distinct phenotype in the schizophrenia spectrum. Figure 4 depicts the position of MNP versus SNP patients in the schizophrenia spectrum.

Intriguingly, the machine learning-derived cluster MNP resembles Kraepelin’s dementia praecox because this phenotype is characterized by a defect (which we have quantified by the G-CoDe and negative symptom scores) coexisting with “productive” or “florid” symptoms (which we have quantified by PHEMFP scores). The MNP phenotype is also reminiscent of the case definition of Eugen Bleuler who conceptualized “schizophrenia” as a psycho-organic disorder (which we have delineated as a neurological disease caused by neuroimmunotoxicity) characterized by primary symptoms (which we have quantified by computing the negative symptom and G-CoDe scores) and accessory symptoms (which we have quantified as psychotic domain scores), such as hallucinations and delusions. Even so, the classification of MNP and SNP based on machine learning does not match Crow’s theory. This is because positive (Type I) or negative (“Type II”) symptoms may be more common, whereas our machine learning shows that these symptoms come from a common core called OSOS.

The distinct category of MNP has a very ambiguous relationship with “deficit schizophrenia”. ^19,20^ On the one hand, we derived the model of MNP using the diagnostic criteria of deficit schizophrenia, with which it exhibits a substantial overlap, ^30,31^ but on the other hand, the machine learning model of MNP differs significantly from the clinical concept of deficit schizophrenia. While the diagnostic criteria of deficit schizophrenia are solely based on negative symptoms, MNP is characterized by more pronounced PHEMFP symptoms, OSOS and single group factor, deficits in the G-CoDe, more severe neuroimmunotoxic pathways, highly specific loss of neuroprotection (natural IgM-mediated autoimmunity, antioxidant levels and antioxidant gene variants), which all distinguish MNP as a diagnostic or nosological subtype which differs qualitatively from SNP and controls. ^30,51,58,64^

Deficit schizophrenia, on the other hand, was conceptualized by Kirkpatrick et al. ^19,20^ as a subtype of schizophrenia defined by persistent “primary” negative symptoms. The diagnostic criteria required the presence of schizophrenia and at least two negative symptoms from a list of six (restricted affect, diminished emotional range, sense of purpose and social drive, poverty of speech, and repression of interests) during clinically stable periods within the previous year. In addition, the negative symptoms must be idiopathic and unrelated to other conditions such as anxiety, depression, drug effects, suspicion, mental retardation, or psychotic symptoms. There are, however, significant issues with the diagnosis of deficit schizophrenia.

First, the relationship between negative symptoms and deficit schizophrenia may be ambiguous, as some authors have concluded that it may be more accurate to view negative symptoms as a dimension rather than a separate disease. ^65^ Additionally, negative and positive dimensions may coexist in the same individual and there is also disagreement regarding whether negative symptoms intensify from a healthy state to schizophrenia with a “fully formed illness” (dimensional theory) or whether type II or deficit schizophrenia is a distinct nosological category (categorical theory). ^31,66^

Second, as described in the previous sections, negative and PHEMFP symptoms (including psychotic symptoms) not only coexist and covary during the stable phase of schizophrenia, but also share a common core that underlies those subdomains. Furthermore, in the stable phase of schizophrenia, negative and PHEMFP symptoms correlate strongly with anxiety and depression to the extent that they all appear to belong to the same latent construct. ^67^ All of these symptom domains are therefore strongly interconnected manifestations of the illness’s symptomatome core, indicating that negative symptoms are not the result of psychosis, depression, or anxiety, but rather are manifestations of the single trait OSOS. Moreover, as indicated by our PLS models, neuroimmunotoxic AOPs explain a large part of the variance in the cognitome and OSOS, as well as the strong covariation between these strongly related components, despite their distinct presentation. Inference shows that negative symptoms should be thought of as a sign of OSOS that is partially determined by AOPs.

Third, Kirkpatrick’s exclusion criteria that negative symptoms may not be caused by depression, anxiety, or psychotic symptoms are nonetheless remarkable. How could one decide whether the negative symptoms which occur in schizophrenia are idiopathic and not the consequence (or cause for that matter) of psychosis, depression or anxiety, which are other features of schizophrenia? Excluding features of an illness to define that interrelated features of the same illness are idiopathic is at least a subjective method. Such subjective criteria have no place in the precision psychiatry approach. Fourth, the MNP patient cluster captures most patients which were diagnosed according to Kirkpatrick’s criteria except that the latter is overinclusive as compared with MNP. ^30^

Fifth, both “schizophrenia” and “deficit” are utterly stigmatizing labels and the label “deficit schizophrenia” does not even describe the disease’s AOP, neurocognitive and symptomatome features. ^30^ Additionally, our machine learning-derived model is conceptually quite different from the clinical case definition of deficit schizophrenia and therefore we labeled the new endophenotype class MNP. ^30^ In fact, this label describes the results of our machine learning results that both negative and PHEMFP symptoms are to a large extent explained by neuroimmunotoxic disorders and cognitive impairments, while the label “major” is used to denote that MNP is the full-fledged qualitatively distinct phenotype as opposed to SNP, which is a less severe phenotype.

In conclusion, our machine learning results demonstrate that a) negative symptoms are not a distinct dimension, as previously described; and b) within schizophrenia a new endophenotype class has been discovered namely MNP, a qualitatively distinct more severe phenotype with increased OSOS and cognitive deficits, and distinct pathophysiology.

## Data Availability

All data produced in the present study are available upon reasonable request to the authors once the data file has been fully exploited by the authors

## Declarations

### Ethics approval and consent to participate

The PLS analyses performed in this paper are based on previously published data. ^51^ Approval was obtained from the Institutional Review Board of the Faculty of Medicine, Chulalongkorn University, Bangkok, Thailand (No 298/57). All participants, as well as the guardians of patients (parents or other close family members) provided written informed consent to take part in the study.

### Consent for publication

Not applicable

### Competing interests

The author declares that he has no competing interests

### Funding

No specific financial support for this sspecific study.

### Authors’ contributions

MM designed the study, performed statistics, and wrote the paper.

## Acknowledgments

none

## Notes

### Competing Interest Statement

The authors have declared no competing interest.

### Author Declarations

The PLS analyses performed in this paper are based on previously published data. 51 Approval was obtained from the Institutional Review Board of the Faculty of Medicine, Chulalongkorn University, Bangkok, Thailand (No 298/57). All participants, as well as the guardians of patients (parents or other close family members) provided written informed consent to take part in the study.

## References

1. GBD 2017 Disease and Injury Incidence and Prevalence Collaborators. Global, regional, and national incidence, prevalence, and years lived with disability for 354 diseases and injuries for 195 countries and territories, 1990-2017: a systematic analysis for the Global Burden of Disease Study 2017. Lancet 2018;392(10159):1789–858. doi: 10.1016/S0140-6736(18)32279-7. Epub 2018 Nov 8. Erratum in: Lancet. 2019 Jun 22;393(10190):e44. PMID: 30496104; PMCID: PMC6227754.

2. Javitt DC. Balancing therapeutic safety and efficacy to improve clinical and economic outcomes in schizophrenia: a clinical overview. Am J Manag Care 2014;20(8 Suppl):S160-5. PMID: 25180705.

3. Jablensky A. Epidemiology of schizophrenia: the global burden of disease and disability. Eur Arch Psychiatry Clin Neurosci 2000;250(6):274–85. doi: 10.1007/s004060070002. PMID: 11153962.

4. Vita A, Barlati S. Recovery from schizophrenia: is it possible? Curr Opin Psychiatry. 2018;31(3):246–55. doi: 10.1097/YCO.0000000000000407. PMID: 29474266.

5. Maes M, Anderson G. False Dogmas in Schizophrenia Research: Toward the Reification of Pathway Phenotypes and Pathway Classes. Front Psychiatry 2021;12:663985. doi: 10.3389/fpsyt.2021.663985. PMID: 34220578; PMCID: PMC8245788.

6. Charlson FJ, Ferrari AJ, Santomauro DF, Diminic S, Stockings E, Scott JG, McGrath JJ, Whiteford HA. Global Epidemiology and Burden of Schizophrenia: Findings From the Global Burden of Disease Study 2016. Schizophr Bull 2018;44(6):1195–1203. doi: 10.1093/schbul/sby058. PMID: 29762765; PMCID: PMC6192504.

7. Hor K, Taylor M. Suicide and schizophrenia: a systematic review of rates and risk factors. J Psychopharmacol 2010;24(4 Suppl):81–90. doi: 10.1177/1359786810385490. PMID: 20923923; PMCID: PMC2951591.

8. Shepherd M. Two faces of Emil Kraepelin. Br J Psychiatry 1995;167(2):174–83. doi: 10.1192/bjp.167.2.174. PMID: 7582666.

9. Engstrom EJ, Weber MM, Burgmair W. Emil Wilhelm Magnus Georg Kraepelin (1856-1926). Am J Psychiatry 2006;163(10):1710. doi: 10.1176/ajp.2006.163.10.1710. PMID: 17012678.

10. Ebert, A., Bar, K. Emil Kraepelin: A pioneer of scientific understanding of psychiatry and psychopharmacology. Ind J Psychiatry 2010;52(2):191–2. doi:http://dx.doi.org/10.4103/0019-5545.64591.

11. Emil Kraepelin. (n.d.). Encyclopedia Britannica Online. Retrieved from http://www.britannica.com/EBchecked/topic/323108/Emil-Kraepelin

12. Berrios, G. E., Luque, R., Villagrán, J. M. Schizophrenia: A conceptual history. Int J Psychol Psychol Therapy 2003;3(2):111–140.

13. Decker HS. How Kraepelinian was Kraepelin? How Kraepelinian are the neo-Kraepelinians?--from Emil Kraepelin to DSM-III. Hist Psychiatry 2007;18(71 Pt 3):337–60. doi: 10.1177/0957154X07078976. PMID: 18175636.

14. Berrios GE. Eugen Bleuler’s place in the history of psychiatry. Schizophr Bull 2011;37(6):1095–8. doi: 10.1093/schbul/sbr132. Epub 2011 Sep 12. PMID: 21914646; PMCID: PMC3196935.

15. Moskowitz A, Heim G. Eugen Bleuler’s Dementia praecox or the group of schizophrenias (1911): a centenary appreciation and reconsideration. Schizophr Bull 2011;37(3):471–9. doi: 10.1093/schbul/sbr016. PMID: 21505113; PMCID: PMC3080676.

16. Jablensky A. The diagnostic concept of schizophrenia: its history, evolution, and future prospects. Dialogues Clin Neurosci 2010;12:271–87.

17. Soares-Weiser K, Maayan N, Bergman H, Davenport C, Kirkham AJ, Grabowski S, Adams CE. First rank symptoms for schizophrenia. Cochrane Database Syst Rev 2015;25:1(1):CD010653. doi: 10.1002/14651858.CD010653.pub2. PMID: 25879096; PMCID: PMC7079421.

18. Crow TJ. Positive and negative schizophrenic symptoms and the role of dopamine. Br J Psychiatry 1980;137:383-6. PMID: 7448479.

19. Kirkpatrick B, Buchanan RW, Ross DE, Carpenter WT Jr. A separate disease within the syndrome of schizophrenia. Arch Gen Psychiatry 2001;58(2):165–71. doi: 10.1001/archpsyc.58.2.165. PMID: 11177118.

20. Kirkpatrick B, Castle D, Murray RM, Carpenter WT Jr. Risk factors for the deficit syndrome of schizophrenia. Schizophr Bull 2000;26(1):233–42. doi: 10.1093/oxfordjournals.schbul.a033443. PMID: 10755684.

21. Kaiser, S., Heekeren, K., Simon, J.J. (2011). The negative symptoms of schizophrenia: category or continuum? Psychopathology 2011;44(6):345–53. doi: 10.1159/000325912. Epub 2011 Aug 17. PMID: 21847001.

22. N.I.H.M. Schizophrenia. As assessed June 14, 2019. https://www.nimh.nih.gov/health/topics/schizophrenia/index.shtml

23. N.H.S. (2019). As assessed June 14, 2019.https://www.nhs.uk/conditions/schizophrenia/symptoms/

24. Maes M, Kanchanatawan B. In (deficit) schizophrenia, a general cognitive decline partly mediates the effects of neuro-immune and neuro-oxidative toxicity on the symptomatome and quality of life. CNS Spectr 2021;12:1–10. doi: 10.1017/S1092852921000419. Epub ahead of print. PMID: 33843548.

25. Ueoka Y, Tomotake M, Tanaka T, Kaneda Y, Taniguchi K, Nakataki M, Numata S, Tayoshi S, Yamauchi K, Sumitani S, Ohmori T, Ueno S, Ohmori T. Quality of life and cognitive dysfunction in people with schizophrenia. Prog Neuropsychopharmacol Biol Psychiatry 2011;35(1):53–9. doi: 10.1016/j.pnpbp.2010.08.018. Epub 2010 Sep 8. PMID: 20804809.

26. Alptekin K, Akvardar Y, Kivircik Akdede BB, Dumlu K, Işik D, Pirinçci F, Yahssin S, Kitiş A. Is quality of life associated with cognitive impairment in schizophrenia? Prog Neuropsychopharmacol Biol Psychiatry 2005;29(2):239–44. doi: 10.1016/j.pnpbp.2004.11.006. Epub 2004 Dec 23. PMID: 15694230.

27. Tolman AW, Kurtz MM. Neurocognitive predictors of objective and subjective quality of life in individuals with schizophrenia: a meta-analytic investigation. Schizophr Bull 2012;38(2):304–15. doi: 10.1093/schbul/sbq077. Epub 2010 Jul 11. PMID: 20624752; PMCID: PMC3283161.

28. Mohamed S, Rosenheck R, Swartz M, Stroup S, Lieberman JA, Keefe RS. Relationship of cognition and psychopathology to functional impairment in schizophrenia. Am J Psychiatry 2008;165(8):978–87. doi: 10.1176/appi.ajp.2008.07111713. Epub 2008 May 1. PMID: 18450928.

29. Keefe RS, Harvey PD. Cognitive impairment in schizophrenia. Handb Exp Pharmacol 2012;(213):11-37. doi: 10.1007/978-3-642-25758-2_2. PMID: 23027411.

30. Kanchanatawan B, Sriswasdi S, Thika S, Stoyanov D, Sirivichayakul S, Carvalho AF, Geffard M, Maes M. Towards a new classification of stable phase schizophrenia into major and simple neuro-cognitive psychosis: Results of unsupervised machine learning analysis. J Eval Clin Pract 2018;24(4):879–91. doi: 10.1111/jep.12945. Epub 2018 May 23. PMID: 29790237.

31. Kanchanatawan B, Sriswasdi S, Thika S, Sirivichayakul S, Carvalho AF, Geffard M, Kubera M, Maes M. Deficit schizophrenia is a discrete diagnostic category defined by neuro-immune and neurocognitive features: results of supervised machine learning. Metab Brain Dis 2018;33(4):1053–67. doi: 10.1007/s11011-018-0208-4. Epub 2018 Mar 11. PMID: 29527624.

32. Al-Hakeim HK, Almulla AF, Al-Dujaili AH, Maes M. Construction of a Neuro-Immune-Cognitive Pathway-Phenotype Underpinning the Phenome of Deficit Schizophrenia. Curr Top Med Chem 2020;20(9):747–58. doi: 10.2174/1568026620666200128143948. PMID: 31994463.

33. Maes M, Vojdani A, Galecki P, Kanchanatawan B. How to Construct a Bottom-Up Nomothetic Network Model and Disclose Novel Nosological Classes by Integrating Risk Resilience and Adverse Outcome Pathways with the Phenome of Schizophrenia. Brain Sci 2020;10(9):645. doi: 10.3390/brainsci10090645. PMID: 32957709; PMCID: PMC7565440.

34. Stoyanov D, Maes MH. How to construct neuroscience-informed psychiatric classification? Towards nomothetic networks psychiatry. World J Psychiatry. 2021;11(1):1–12. doi: 10.5498/wjp.v11.i1.1. PMID: 33511042; PMCID: PMC7805251.

35. Ringle, C.M.; da Silva, D.; Bido, D. Structural equation modeling with the SmartPLS. Brazilian Journal of Marketing - BJM Revista Brasileira de Marketing – ReMark Edição Especial 2014,13,n2.

36. Hair, J. F., Hult, G. T. M., Ringle, C. M., and Sarstedt, M. A Primer on Partial Least Squares Structural Equation Modeling (PLS-SEM), 3rd Ed., Thousand Oakes, CA: Sage, 2022.

37. Rigdon, E. E. Rethinking partial least squares path modeling: In praise of simple methods. Long Range Planning, 2012;45(5–6):341–58.

38. Henseler, J., Ringle, C.M. & Sarstedt, M. A new criterion for assessing discriminant validity in variance-based structural equation modeling. J Acad Mark Sci 2015;43:115–135 https://doi.org/10.1007/s11747-014-0403-8

39. Gefen, D., & Straub, D. W. A practical guide to factorial validity using PLS-Graph: tutorial and annotated example. Communications of the AIS 2005;16:91–109

40. Chin, W. W. How to write up and report PLS analyses. In V. Esposito Vinzi, W. W. Chin, J. Henseler, & H. Wang (Eds.), Handbook of partial least squares: concepts, methods and applications in marketing and related fields, 2010, pp. 655–690, Berlin: Springer.

41. Kline, R.B. Principles and practice of structural equation modeling. New York, Guilford Press, 2011.

42. Teo, T. Factors influencing teachers’ intention to use technology: Model development and test. Computers & Education 2011;57:2432–40.

43. Andreasen NC. The Scale for the Assessment of Negative Symptoms (SANS): conceptual and theoretical foundations. Br J Psychiatry Suppl 1989;(7):49-58. PMID: 2695141.

44. Eearly Psychosis Intervention. Symptoms of psychosis. Symptoms of Psychosis - Early Psychosis Intervention. As accessed 26-5-2022

45. Noto C, Ota VK, Santoro ML, Gouvea ES, Silva PN, Spindola LM, Cordeiro Q, Bressan RA, Gadelha A, Brietzke E, Belangero SI, Maes M. Depression, Cytokine, and Cytokine by Treatment Interactions Modulate Gene Expression in Antipsychotic Naïve First Episode Psychosis. Mol Neurobiol 2016;53(8):5701–9. doi: 10.1007/s12035-015-9489-3. Epub 2015 Oct 22. PMID: 26491028.

46. Kanchanatawan B, Thika S, Sirivichayakul S, Carvalho AF, Geffard M, Maes M. In Schizophrenia, Depression, Anxiety, and Physiosomatic Symptoms Are Strongly Related to Psychotic Symptoms and Excitation, Impairments in Episodic Memory, and Increased Production of Neurotoxic Tryptophan Catabolites: a Multivariate and Machine Learning Study. Neurotox Res 2018;33(3):641–55. doi: 10.1007/s12640-018-9868-4. Epub 2018 Jan 29. PMID: 29380275.

47. Kay SR, Fiszbein A, Opler LA. The positive and negative syndrome scale (PANSS) for schizophrenia. Schizophr Bull. 1987;13(2):261–76. doi: 10.1093/schbul/13.2.261. PMID: 3616518.

48. Overall, J.E.; Gorham, D.R. The brief psychiatric rating scale. Psycholog Rep 1962;10:799–812.

49. Sirivichayakul S, Kanchanatawan B, Thika S, Carvalho AF, Maes M. Eotaxin, an Endogenous Cognitive Deteriorating Chemokine (ECDC), Is a Major Contributor to Cognitive Decline in Normal People and to Executive, Memory, and Sustained Attention Deficits, Formal Thought Disorders, and Psychopathology in Schizophrenia Patients. Neurotox Res 2019;35(1):122–38. doi: 10.1007/s12640-018-9937-8. Epub 2018 Jul 28. PMID: 30056534.

50. Maes M, Sirivichayakul S, Kanchanatawan B, Carvalho AF. In schizophrenia, psychomotor retardation is associated with executive and memory impairments, negative and psychotic symptoms, neurotoxic immune products and lower natural IgM to malondialdehyde. World J Biol Psychiatry 2020;21(5):383–401. doi: 10.1080/15622975.2019.1701203. Epub 2020 Feb 7. PMID: 32031479.

51. Maes M, Vojdani A, Geffard M, Moreira EG, Barbosa DS, Michelin AP, Semeão LO, Sirivichayakul S, Kanchanatawan B. Schizophrenia phenomenology comprises a bifactorial general severity and a single-group factor, which are differently associated with neurotoxic immune and immune-regulatory pathways. Biomol Concepts 2019;10(1):209–25. doi: 10.1515/bmc-2019-0023. PMID: 31734647.

52. Hamilton M. A rating scale for depression. J Neurol Neurosurg Psychiatry 1960;23(1):56–62. doi: 10.1136/jnnp.23.1.56. PMID: 14399272; PMCID: PMC495331.

53. CANTAB. The most validated cognitive research software. http://www.cambridgecognition.com/cantab/ October 1, 2018.

54. Bland JM, Altman DG. Correlation in restricted ranges of data. BMJ 2011;11:342:d556. doi: 10.1136/bmj.d556. PMID: 21398359.

55. Karsner D. Restriction of Range: Definition & Examples. Study.com Restriction of Range: Definition & Examples - Video & Lesson Transcript | Study.com. As accessed 26-5-2022.

56. Sackett PR, Yang H. Correction for range restriction: an expanded typology. J Appl Psychol 2000;85(1):112–8. doi: 10.1037/0021-9010.85.1.112. PMID: 10740961.

57. Maes M, Vojdani A, Sirivichayakul S, Barbosa DS, Kanchanatawan B. Inflammatory and Oxidative Pathways Are New Drug Targets in Multiple Episode Schizophrenia and Leaky Gut, Klebsiella pneumoniae, and C1q Immune Complexes Are Additional Drug Targets in First Episode Schizophrenia. Mol Neurobiol 2021;58(7):3319–34. doi: 10.1007/s12035-021-02343-8. Epub 2021 Mar 6. PMID: 33675500.

58. Almulla AF, Al-Hakeim HK, Maes M. Schizophrenia phenomenology revisited: positive and negative symptoms are strongly related reflective manifestations of an underlying single trait indicating overall severity of schizophrenia. CNS Spectr 2021;26(4):368–77. doi: 10.1017/S1092852920001182. Epub 2020 May 20. PMID: 32431263.

59. Orellana G, Slachevsky A. Executive functioning in schizophrenia. Front Psychiatry 2013;24;4:35. doi: 10.3389/fpsyt.2013.00035. PMID: 23805107; PMCID: PMC3690455.

60. Corlett PR, Honey GD, Fletcher PC. From prediction error to psychosis: ketamine as a pharmacological model of delusions. J Psychopharmacol 2007;21(3):238–52. doi: 10.1177/0269881107077716. PMID: 17591652.

61. Brewer WJ, Francey SM, Wood SJ, Jackson HJ, Pantelis C, Phillips LJ, Yung AR, Anderson VA, McGorry PD. Memory impairments identified in people at ultra-high risk for psychosis who later develop first-episode psychosis. Am J Psychiatry 2005;162(1):71–8. doi: 10.1176/appi.ajp.162.1.71. PMID: 15625204.

62. Hawkins KA, Addington J, Keefe RS, Christensen B, Perkins DO, Zipurksy R, Woods SW, Miller TJ, Marquez E, Breier A, McGlashan TH. Neuropsychological status of subjects at high risk for a first episode of psychosis. Schizophr Res 2004;67(2-3):115-22. doi: 10.1016/j.schres.2003.08.007. PMID: 14984870.

63. Fillenbaum GG, van Belle G, Morris JC, Mohs RC, Mirra SS, Davis PC, Tariot PN, Silverman JM, Clark CM, Welsh-Bohmer KA, Heyman A. Consortium to Establish a Registry for Alzheimer’s Disease (CERAD): the first twenty years. Alzheimers Dement 2008;4(2):96–109. doi: 10.1016/j.jalz.2007.08.005. PMID: 18631955; PMCID: PMC2808763.

64. Al-Hakeim HK, Almulla AF, Maes M. The Neuroimmune and Neurotoxic Fingerprint of Major Neurocognitive Psychosis or Deficit Schizophrenia: a Supervised Machine Learning Study. Neurotox Res 2020;37(3):753–71. doi: 10.1007/s12640-019-00112-z. Epub 2020 Jan 8. PMID: 31916129

65. Roy MA, DeVriendt X. Symptômes positifs et négatifs de la schizophrénie: une mise à jour [Positive and negative symptoms in schizophrenia: a current overview]. Can J Psychiatry 1994;39(7):407-14. French. doi: 10.1177/070674379403900704. PMID: 7987782.

66. Takahashi S. Heterogeneity of schizophrenia: Genetic and symptomatic factors. Am J Med Genet B Neuropsychiatr Genet 2013;162B:648–652.

67. Almulla AF, Al-Rawi KF, Maes M, Al-Hakeim HK. In schizophrenia, immune-inflammatory pathways are strongly associated with depressive and anxiety symptoms, which are part of a latent trait which comprises neurocognitive impairments and schizophrenia symptoms. J Affect Disord 2021;287:316–26. doi: 10.1016/j.jad.2021.03.062. Epub 2021 Mar 26. PMID: 33812245.

